# Impact of COVID-19 on College Students’ at One of the Most Diverse Campuses in the United States: A Factor Analysis of Survey Data

**DOI:** 10.1101/2022.06.14.22276416

**Authors:** Bowen Liu, Edward Huynh, Chengcheng Li, Qing Wu

**Affiliations:** Nevada Institute of Personalized Medicine, College of Science, University of Nevada Las Vegas, Las Vegas, Nevada, USA; Department of Mathematical Sciences, University of Nevada, Las Vegas, Las Vegas, NV, USA; Department of Epidemiology and Biostatistics, School of Public Health, University of Nevada Las Vegas, Las Vegas, Nevada, USA; The Open University of China, No. 75 Fuxing Rd, Haidian District, Beijing, 100039

**Keywords:** Survey Study, Mental Health, Exploratory Factor Analysis, COVID-19, Perceived Stress

## Abstract

**Objective:** This survey study is designed to understand the impact of the COVID-19 pandemic on stress among specific sub-populations of college students.

**Design, Settings and Participants:** An online questionnaire was sent to the students from University of Nevada, Las Vegas between 2020 October and December to assess the psychological impact of COVID-19. A total of 2,091 respondents signed the consent form online and their responses were collected.

**Methods:** The Perceived Stress levels of college students were analyzed via exploratory factor analysis of a survey of college students, which was collected at the University of Nevada, Las Vegas (UNLV). An explanatory factor analysis was carried out on the Perceived Stress Scale (PSS-10) results. We subsequently analyzed each factor using stepwise linear regression that focused on various socio-demographic groups.

**Results:** A two-factor model was obtained using the explanatory factor analysis. After comparing with the past studies that investigated the factor structure of the PSS-10 scale, we identified these two factors as “Anxiety” and “Irritability”. The subsequent stepwise linear regression analysis suggested that gender and age (*P*< 0.01) are significantly associated with both factors. However, the ethnicities of students are not significantly associated with both factors.

**Conclusions:** To our knowledge, this is the first study that assessed the perceived stress of university students in the US during the COVID-19 pandemic. We showed that the PSS-10 scale could be summarized as a two-factor structure through an exploratory factor analysis. A stepwise regression approach was used and we found both of the factors are significantly associated with the gender of the participants. However, we found no significant association between both factors and ethnicity. In summary, our findings will help identify students with higher risk for stress and mental health issues in pandemics and future crises.

**Strengths and limitations of this study:**

➣ To our knowledge, this is the first study that assessed the perceived stress of university students with PSS-10 scale in the US during the COVID-19 pandemic.
➣ Ethnicities of the participants are not significantly associated with the perceived stress. This finding is different compared to most of the previous studies.
➣ Only quantitative questions were used in this survey study. To gain a better understanding for the psychological impact of COVID-19 on students, qualitative questions need to be included in the future survey studies.
➣ The fact that only the participants who completed the PSS-10 scale were included in the quantitative analysis in this study leads to potential selection bias.
➣ The proportions of African American students and Pacific Islander students are low within the survey sample compared to the demographics of UNLV. This leads to potential volunteer bias.

## Introduction

The United States has reported more than 500,000 deaths due to the COVID-19 pandemic, with over 49 million total reported cases of COVID-19 as of December 2021^1^. Studies have shown a significant effect on students’ mental health, including anxiety and depression, resulting from the onset of the COVID-19 pandemic ^2–4^. Due to the transition to online models of instruction, many university campuses have closed, and resident students are forced to live away from campus ^5^. Furthermore, the unexpected shift from in-person classes to online instruction has proven difficult for students who do not have free or easy access to digital resources ^5^. In addition, increased levels of anxiety and depression have been more prevalently observed within specific ethnic communities and have been especially difficult for women and for those of Chinese descent ^6–8^.

Many survey results have been collected and analyzed regarding the impact of COVID-19 concerning mental health; however, those have been mainly limited to studies in developing countries ^9^. In most of the studies focusing on college students within the United States, anxiety and depression were measured using Patient Health Questionnaire Depression scale (PHQ-8) and the Generalized Anxiety Disorder scale (GAD-7) ^3,10,11^. Some have also included deeper analyses regarding the impact of race and ethnicity with respect to increased levels of stress, anxiety, and depression ^12^. On the other hand, very few studies investigated the mental health of female college students during the pandemic ^13,14^. Moreover, we discovered that there had been only one study that was conducted for the University of Nevada, Las Vegas (UNLV) student population, which is considered the most diverse student body in the United States ^15^. However, that study only considered changes in depression and physical activity, whereas our work focuses directly on stress.

The objective of this study was to conduct a survey-based assessment of stress among college students at UNLV ^16^ during the COVID-19 pandemic. We measured stress levels among college students by using the Perceived Stress Scale (PSS-10). We sought to identify severity levels of stress related to COVID-19, validate the factors under the PPS-10, and examine the relationships between those factors and the demographic variables (gender, age, financial situation, marital status, class standing, employment status, and ethnicity).

## Methods

### Ethics

This study was approved by the UNLV Institutional Review Board (IRB) in August 2020. All participants signed an informed consent form that the IRB had approved.

### Recruitment of the Participants

Student email addresses were requested by the Principal Investigator (PI) of this study, using the inclusion criteria of currently enrolled UNLV students, age 18 and older, including undergraduate, graduate, professional, and/or non-degree seeking students provided from the Registrar’s Office at UNLV. A recruitment email was sent to each qualifying participant, with a link generated by the Qualtrics online survey platform, to invite them to voluntarily participate in the study. Informed consent was indicated once the willing participants clicked the survey link in the recruitment email. Only consenting participants were directed to respond to the survey.

### Measures

PSS-10 (Perceived Stress Scale) was used to measure the perceived stress. PSS-10 includes 10 questions and the participants of this study choose their degree of agreement (4 = Very often; 3 = Fairly often; 2 = Sometimes; 1 = Almost Never; 0 = Never) ^17^. The scale items measure stress and the ability to cope with the stress. The range of the PSS-10 scale is 0-40. A higher PSS-10 score indicates a higher level of stress. We provided the PSS-10 scale as one of the supplementary materials (**Supplementary File 1**). Since the PSS-10 scale is not a diagnostic tool, there is no pre-specified threshold to classify the level of stress. However, several previous studies used PSS-10 scores of 0-13, 14-26, and 27-40 to categorize low, moderate, and high-stress levels, correspondingly^18–21^. A previous study showed that PSS-10 has good reliability measures among college students ^22^. Demographic information was also collected, including gender, age, ethnicity, class standing (i.e., Undergraduate [1^st^ year, 2^nd^ year, 3^rd^ year, 4^th^ year, 5^th^ year or more], Graduate [Master’s, Doctorate] or Non-Degree Seeking), marital status (i.e., Married, Widowed, Divorced, Separated, Partnered, Single or Other), and financial situation.

### Statistical Analyses

The data were downloaded from the Qualtrics online survey platform. Statistical analysis was conducted using R statistical software. Univariate analysis of students’ stress from the COVID-19 pandemic was done on the responses from the participants who completed all 10 questions from PSS-10. The Kruskal-Wallis test and the one-way ANOVA F-test were used to assess the relationship between the student’s stress and the demographic variables. Furthermore, a factor analysis using the principal axis and the varimax rotation methods was conducted on the responses. The Kaiser-Meyer-Olkin (KMO) and Bartlett’s Tests were used to check for factorization and validate variance homogeneity. Based on the previous factor analysis on PSS-10 ^22,23^, we extracted the factors correspondingly in our exploratory factor analysis. Stepwise linear regression using Akaike Information Criterion was applied to assess the association between demographic variables and the factors extracted in the factor analysis stage.

### Patient and Public Involvement

The participants of this study were students at UNLV during 2020 fall semester. A major aim of the study is to give the students an opportunity to express their own feelings against a global pandemic like COVID-19. We sent recruitment emails to the UNLV students for their contributions to the survey, with the permission from UNLV Registrar’s Office. The research question of this study was to provide an assessment of mental stress among college students during COVID-19. We designed the study based on the previous and ongoing survey studies within different universities with our own understandings about how to efficiently involve UNLV students in our study. The study was completely open during all the steps of this study and we made the student aware of the possible time taken to finish the survey via Qualtrics online survey platform.

## Results

### Sample Demographics

A total of 2,091 responses were collected via Qualtrics. After removing the responses which did not complete all 10 questions from PSS-10, 1,699 responses remained for analysis. Among these 1,699 students, 1,152 (67.8%) were females. The sample includes both undergraduate (n = 1,303, 76.7%) and graduate students (n = 364, 22.4%). Thirty eight point three percent of the students (n = 650) identified themselves as “White/Caucasian,” 23.7% (n = 403) were “Hispanic/Latino,” 20.1% were (n = 341) are “Asian/Asian American,” 6.8% (n = 115) classified themselves as “Biracial/Multiracial”, 5.7% (n = 96) were “Black/African American.” In addition, 1.9% (n = 32) identified themselves as Pacific Islanders/Native Hawaiian, 1.5% identified themselves as “Middle Eastern or Northern African (MENA)/Arabic Origin,” and 0.9% (n = 16) identified themselves as “American Indian/Native Alaskan.” Age, gender, marital status, ethnicity, class standing, employment status, and financial situation are summarized in **Table 1**.

**Table 1:**
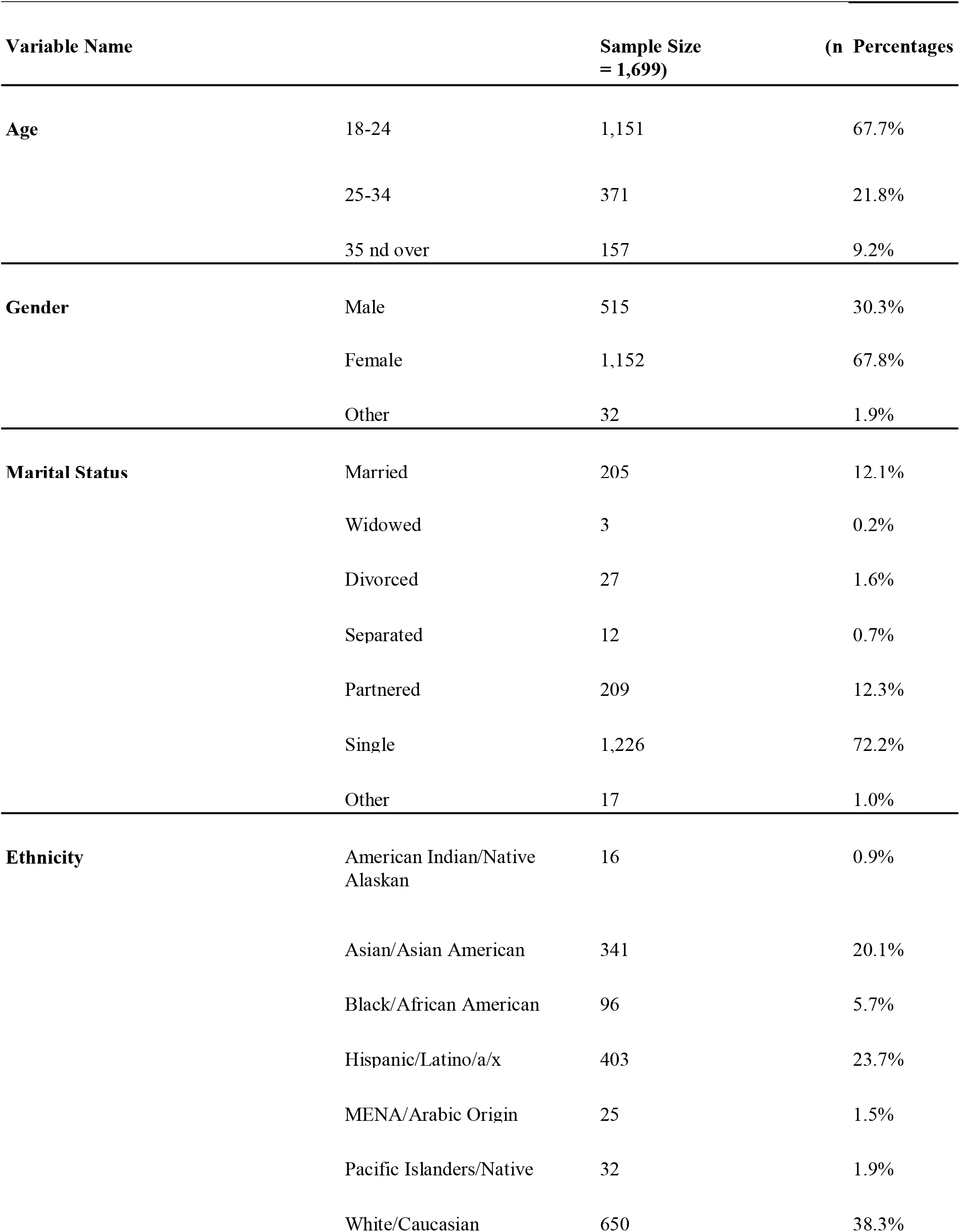
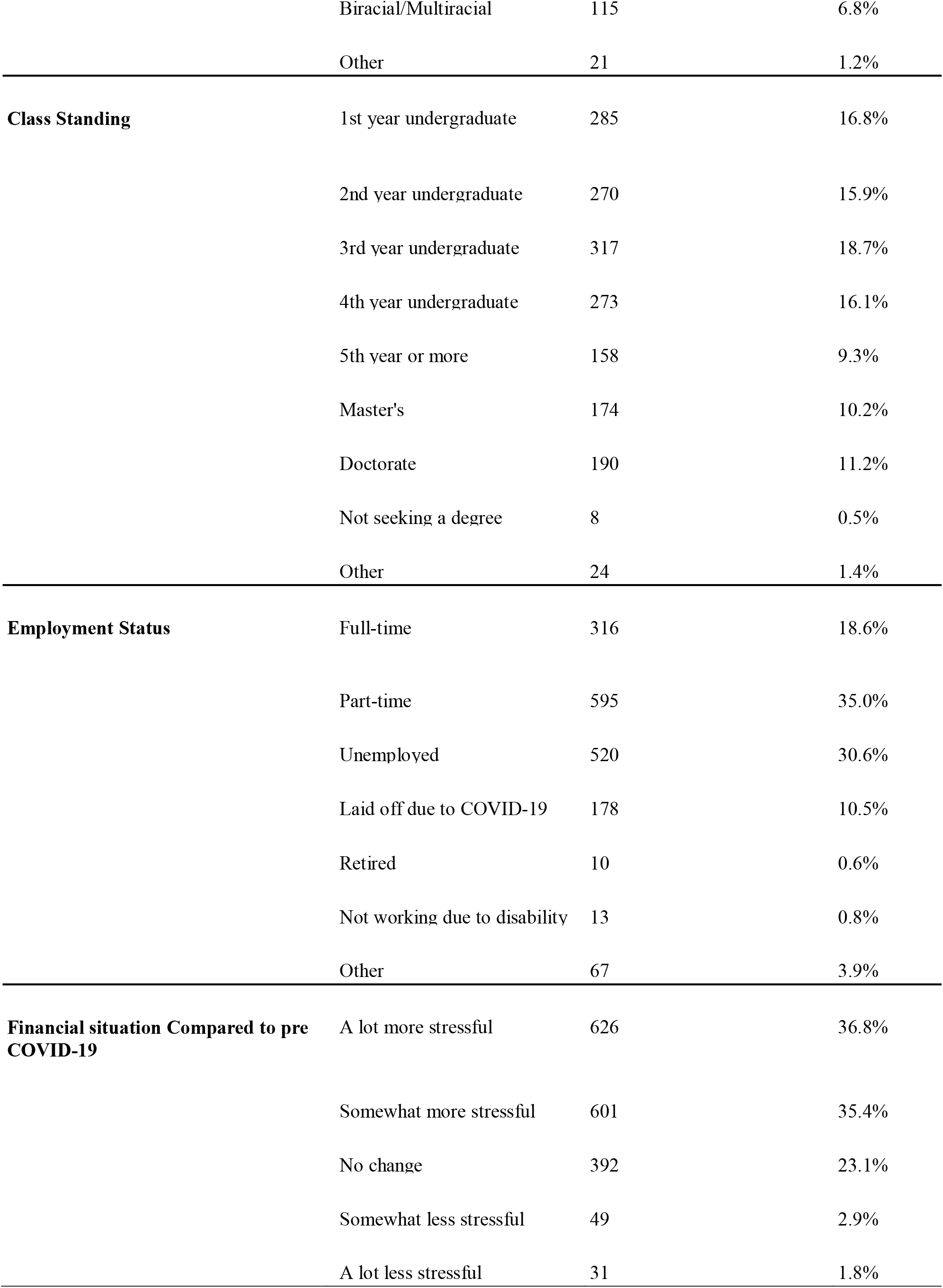
Demographic Information

| Variable Name |  | Sample Size<br>= 1,699) | (n Percentages |
| --- | --- | --- | --- |
| Age | 18-24 | 1,151 | 67.7% |
|  | 25-34 | 371 | 21.8% |
|  | 35 and over | 157 | 9.2% |
| Gender | Male | 515 | 30.3% |
|  | Female | 1,152 | 67.8% |
|  | Other | 32 | 1.9% |
| Marital Status | Married | 205 | 12.1% |
|  | Widowed | 3 | 0.2% |
|  | Divorced | 27 | 1.6% |
|  | Separated | 12 | 0.7% |
|  | Partnered | 209 | 12.3% |
|  | Single | 1,226 | 72.2% |
|  | Other | 17 | 1.0% |
| Ethnicity | American Indian/Native Alaskan | 16 | 0.9% |
|  | Asian/Asian American | 341 | 20.1% |
|  | Black/African American | 96 | 5.7% |
|  | Hispanic/Latino/a/x | 403 | 23.7% |
|  | MENA/Arabic Origin | 25 | 1.5% |
|  | Pacific Islanders/Native | 32 | 1.9% |
|  | White/Caucasian | 650 | 38.3% |
|  | Biracial/Multiracial | 115 | 6.8% |
|  | Other | 21 | 1.2% |
| **Class Standing** | 1st year undergraduate | 285 | 16.8% |
|  | 2nd year undergraduate | 270 | 15.9% |
|  | 3rd year undergraduate | 317 | 18.7% |
|  | 4th year undergraduate | 273 | 16.1% |
|  | 5th year or more | 158 | 9.3% |
|  | Master's | 174 | 10.2% |
|  | Doctorate | 190 | 11.2% |
|  | Not seeking a degree | 8 | 0.5% |
|  | Other | 24 | 1.4% |
| **Employment Status** | Full-time | 316 | 18.6% |
|  | Part-time | 595 | 35.0% |
|  | Unemployed | 520 | 30.6% |
|  | Laid off due to COVID-19 | 178 | 10.5% |
|  | Retired | 10 | 0.6% |
|  | Not working due to disability | 13 | 0.8% |
|  | Other | 67 | 3.9% |
| **Financial situation Compared to pre COVID-19** | A lot more stressful | 626 | 36.8% |
|  | Somewhat more stressful | 601 | 35.4% |
|  | No change | 392 | 23.1% |
|  | Somewhat less stressful | 49 | 2.9% |
|  | A lot less stressful | 31 | 1.8% |

### Descriptive Statistics

For all the participants who completed PSS-10, the average PSS score was 21.55. Eighty-three-point nine percent of these participants perceived moderate to severe stress. The mean PSS scores for different groups are listed in **Table 2**. The female participants had a mean PSS score of 22.20, while the male participants had a mean PSS score of 19.7. The mean PSS score of the participants who reported their gender as “other” was 27.4. Among all the age groups, the participants between 18 and 24 had the highest PSS score at 22.2, while the participants 35 years or over had the lowest PSS score at 18.8. Among all the ethnic groups, the African American/Black participants had the lowest mean PSS score, at 20.3, and the MENA/Arabic origin participants had the highest mean PSS score, at 22.3. Among all the class standings, the undergraduate participants generally had higher mean PSS scores (Freshmen: 20.7; Sophomore: 22.5; Junior: 22.9; Senior: 22.3; 5th year or more: 22.5) compared to all other students (Master’s: 19.6; Ph.D.: 19.5; Non-degree seeking: 19.4). The participants who identified themselves as “a lot more stressful” in financial situations during COVID-19 had the highest mean PSS score, at 24.7. In contrast, the participants who identified themselves as “a lot less stressful” in financial situations had the lowest mean PSS score, at 17.1. For the employment status, the retired participants had the lowest mean PSS score, at 16.6, and the participants who were not working due to disability had the highest mean PSS score, at 26.5. The participants who lost their jobs due to COVID-19 also had a high mean PSS score, at 23.9. The married students had the lowest mean PSS score, 19.0, while the separated students had the highest mean PSS score, 26.4.

**Table 2:** Univariate Analysis of UNLV Students’ Stress about the COVID-19 pandemic

| Variable |  | Total | Low Stress (%) | Moderate Stress (%) | High Stress (%) | Mean PSS-10 Score | One-Way ANOVA<sup>1</sup> | Kruskal-Wallis<sup>2</sup> |
| --- | --- | --- | --- | --- | --- | --- | --- | --- |
| Age | 18-24 | 1151 | 153 (13.3%) | 661 (19.1%) | 337 (29.3%) | 22.2 | <0.01 | <0.01 |
|  | 25-34 | 371 | 71 (19.1%) | 204 (55.0%) | 96 (25.9%) | 20.8 |  |  |
|  | 35 and over | 157 | 49 (23.3%) | 93 (21.7%) | 35 (55.0%) | 18.8 |  |  |
| Gender | Male | 515 | 117 (22.7%) | 292 (56.7%) | 106 (20.6%) | 19.7 | <0.01 | <0.01 |
|  | Female | 1152 | 154 (13.4%) | 654 (56.8%) | 344 (29.9%) | 22.2 |  |  |
|  | Other | 32 | 2 (6.2%) | 12 (37.5%) | 18 (56.3%) | 27.4 |  |  |
| Marital Status | Married | 205 | 54 (26.3%) | 113 (55.1%) | 38 (18.5%) | 19.0 | <0.01 | <0.01 |
|  | Widowed | 3 | 0 (0%) | 3 (100%) | 0 (0%) | 22.3 |  |  |
|  | Divorced | 27 | 6 (22.2%) | 13 (48.1%) | 8 (29.6%) | 20.6 |  |  |
|  | Separated | 12 | 1 (8.3%) | 5 (41.7%) | 6 (50.0%) | 26.4 |  |  |
|  | Partnered | 209 | 22 (10.7%) | 116 (56.3%) | 71 (34.5%) | 22.7 |  |  |
|  | Single | 1226 | 186 (15.2%) | 699 (57.0%) | 341 (27.8%) | 21.7 |  |  |
|  | Other | 17 | 4 (23.5%) | 9 (53.0%) | 4 (23.5%) | 20.5 |  |  |
| Ethnicity | American Indian/Native Alaskan | 16 | 3 (18.7%) | 8 (50.0%) | 5 (31.3%) | 21.8 | 0.36 | 0.45 |
|  | Asian/Asian American | 341 | 52 (15.2%) | 212 (62.2%) | 77 (22.6%) | 21.1 |  |  |
|  | Black/African American | 96 | 21 (21.9%) | 48 (50.0%) | 27 (28.1%) | 20.3 |  |  |
|  | Hispanic/Latino/a/x | 403 | 50 (12.4%) | 240 (59.6%) | 113 (28.0%) | 22.2 |  |  |
|  | MENA/Arabic Origin | 25 | 4 (16.0%) | 13 (52.0%) | 8 (32.0%) | 22.3 |  |  |
|  | Pacific Islanders/Native Hawaiian | 32 | 6 (18.7%) | 19 (59.4%) | 7 (21.9%) | 21.3 |  |  |
|  | White/Caucasian | 650 | 117 (18.0%) | 342 (52.6%) | 191 (29.4%) | 21.5 |  |  |
|  | Biracial/Multiracial | 115 | 14 (12.2%) | 67 (58.3%) | 34 (29.6%) | 21.9 |  |  |
|  | Other | 21 | 6 (28.6%) | 9 (42.8%) | 6 (28.6%) | 19.6 |  |  |
| **Class Standing** | 1st year undergraduate | 285 | 45 (15.8%) | 182 (63.9%) | 58 (20.3%) | 20.6 | <0.01 | <0.01 |
|  | 2nd year undergraduate | 270 | 28 (10.4%) | 161 (59.6%) | 81 (30.0%) | 22.5 |  |  |
|  | 3rd year undergraduate | 317 | 40 (12.6%) | 165 (52.1%) | 112 (35.3%) | 22.9 |  |  |
|  | 4th year undergraduate | 273 | 45 (16.5%) | 141 (51.6%) | 87 (31.9%) | 22.3 |  |  |
|  | 5th year or more undergraduate | 158 | 23 (14.5%) | 84 (53.2%) | 51 (32.3%) | 22.5 |  |  |
|  | Master's | 174 | 40 (23.0%) | 100 (57.5%) | 34 (19.5%) | 19.6 |  |  |
|  | Doctorate | 190 | 46 (24.2%) | 106 (55.8%) | 38 (20.0%) | 19.5 |  |  |
|  | Not seeking a degree | 8 | 0 (0%) | 7 (87.5%) | 1 (12.5%) | 19.4 |  |  |
|  | Other | 24 | 6 (25.0%) | 12 (50.0%) | 6 (25.0%) | 20.8 |  |  |
| **Employment Status** | Full-time | 316 | 60 (19.0%) | 183 (57.9%) | 73 (23.1%) | 20.8 | <0.01 | <0.01 |
|  | Part-time | 595 | 89 (15.0%) | 336 (56.5%) | 170 (28.6%) | 21.7 |  |  |
|  | Unemployed | 520 | 87 (16.7%) | 306 (58.8%) | 127 (24.4%) | 21.1 |  |  |
|  | Laid off due to COVID-19 | 178 | 18 (10.1%) | 85 (47.8%) | 75 (42.1%) | 23.9 |  |  |
|  | Retired | 10 | 4 (40.0%) | 4 (40.0%) | 2 (20.0%) | 16.6 |  |  |
|  | Not working due to disability | 13 | 0 (0%) | 6 (46.2%) | 7 (53.8%) | 26.5 |  |  |
|  | Other | 67 | 15 (22.4%) | 38 (56.7%) | 14 (20.9%) | 20.3 |  |  |
| **Financial situation Compared to pre COVID-19** | A lot of more stressful | 626 | 41 (6.5%) | 320 (51.1%) | 265 (42.3%) | 24.7 | <0.01 | <0.01 |
|  | Somewhat more stressful | 601 | 95 (15.8%) | 375 (62.4%) | 131 (21.8%) | 21.0 |  |  |
|  | No change | 392 | 117 (29.8%) | 220 (56.1%) | 55 (14.0%) | 17.9 |  |  |
|  | Somewhat less stressful | 49 | 9 (18.4%) | 28 (57.1%) | 12 (24.4%) | 20.2 |  |  |
|  | A lot less stressful | 31 | 11 (35.5%) | 15 (48.4%) | 5 (16.1%) | 17.1 |  |  |

<sup>1</sup> The p-values are obtained via F tests in one-way ANOVA. The F tests were performed to test the equality of the means for the groups within each variable.
<sup>2</sup> The p-values are obtained via Kruskal-Wallis tests. The Kruskal-Wallis tests were performed to test the equality of the medians for the groups within each variable.

### Univariate Analysis

The relationship between students’ stress and the demographic variables is also presented in Table 2. The PSS scores for the students were calculated following the instructions from the PSS-10 scale ^17^. For both of the statistical tests, variables including age (*P*<0.01), gender (*P*<0.01), marital status (*P*<0.01), employment status (*P*<0.01), class standing (*P*<0.01), and financial situation (*P*<0.01) demonstrated significant effects on stress (**Table 2**). However, ethnicity did not significantly influence students’ stress (**Table 2**).

### Factor Analysis

An initial factor analysis was performed on all ten measures from the PSS-10. The Kaiser-Meyer-Olkin (KMO) value of 0.90 justified that the sample was factorable. Bartlett’s test was performed to confirm the homogeneity of variance (χ^2^(45) = 8276.4, ***P*** < 0.01). We obtained the anti-image correlation matrix to determine if any ten items should be dropped. There were five items within the anti-image correlation matrix with the corresponding diagonal elements < 0.5 (Q1, Q2, Q3, Q6, Q10). These items were not included in the final step of factor analysis.

The final step of the factor analysis was conducted using five items (Q4, Q5, Q7, Q8, Q9). KMO value of 0.81 indicates the sample was factorable, and Bartlett’s test provided significant evidence for homogeneity of variance (χ^2^(10) = 2215.8, ***P*** < 0.001). Communalities were above 0.40 except for Q9. However, we did not drop the item since it still has a communality over 0.2. We found it necessary to keep it in the next step of the factor analysis in order to more accurately interpret the model.

We extracted two factors after we inspected the scree plot (**Supplementary Figure 1**). The two factors correspondingly explained 52.7% and 17.5% of the total variance. The cumulative percentage of variance explained by these two factors was 70.2%. The rotated component matrix and the communalities for five measures are provided in **Table 3**. After comparing the factor analysis result with the past literature on PSS-10 ^23,24^, we named the first-factor “Irritability” and the second-factor “Anxiety.” The factor “Anxiety” contained only one item. However, we kept it because this factor provided a different aspect on perceived stress when compared to the factor of “Irritability.”

**Table 3:** Factor Analysis Summary with Rotated Component Matrix*

|  | Factor1 (Irritability) | Factor2 (Anxiety) | Communalities | Diagonal of anti-image matrix |
| --- | --- | --- | --- | --- |
| PSS_4 | 0.727 | 0.199 | 0.569 | 0.633 |
| PSS_5 | 0.568 | 0.419 | 0.498 | 0.567 |
| PSS_7 | 0.491 | 0.414 | 0.412 | 0.634 |
| PSS_8 | 0.555 | 0.524 | 0.583 | 0.507 |
| PSS_9 | 0.155 | 0.468 | 0.243 | 0.567 |

\* Varimax rotation with Principal Axis Factoring was used. The objects with corresponding diagonal elements > 0.5 in the anti-image matrix were kept.

We constructed the bar plots of mean factor scores with respect to different categories within demographic variables. **Figure 1** shows that the students who identified their gender as “other” have higher irritability and anxiety scores compared to the male and female students. The students in a much more stressful financial situation during COVID also have higher irritability and anxiety scores compared to other students (**Figure 2**). The students of Middle East Origin experienced higher irritability scores compared to other students, and the African American students had lower anxiety scores compared to other students **(Figure 3)**. The plots of mean factor scores with respect to class standing **(Supplementary Figure 2)** and marital status **(Supplementary Figure 3)** are provided in the supplementary.

**Figure 1:**
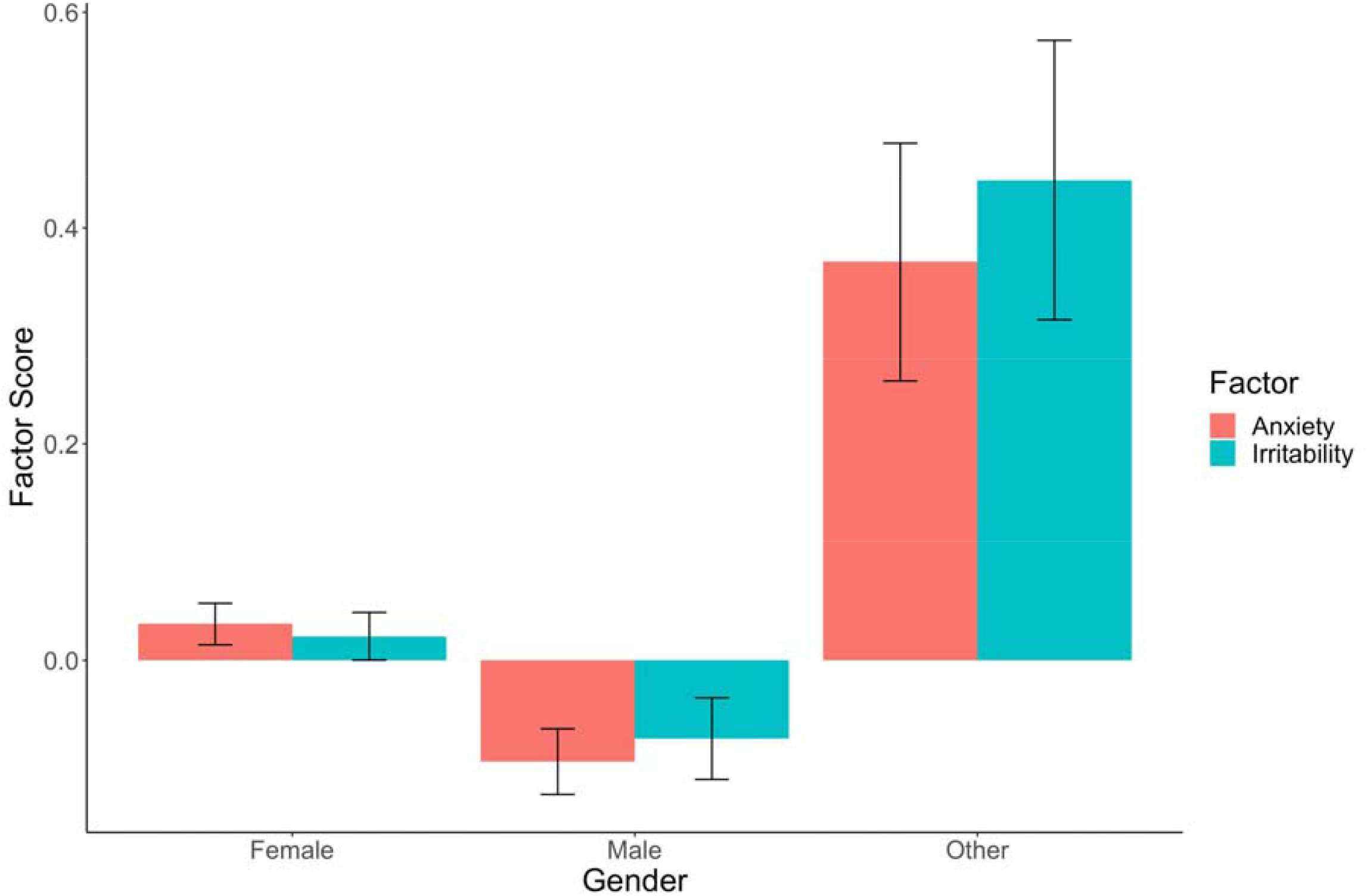
Mean Factor Scores for UNLV Students (n = 1,699) by Gender. Means and standard errors are presented.

**Figure 2:**
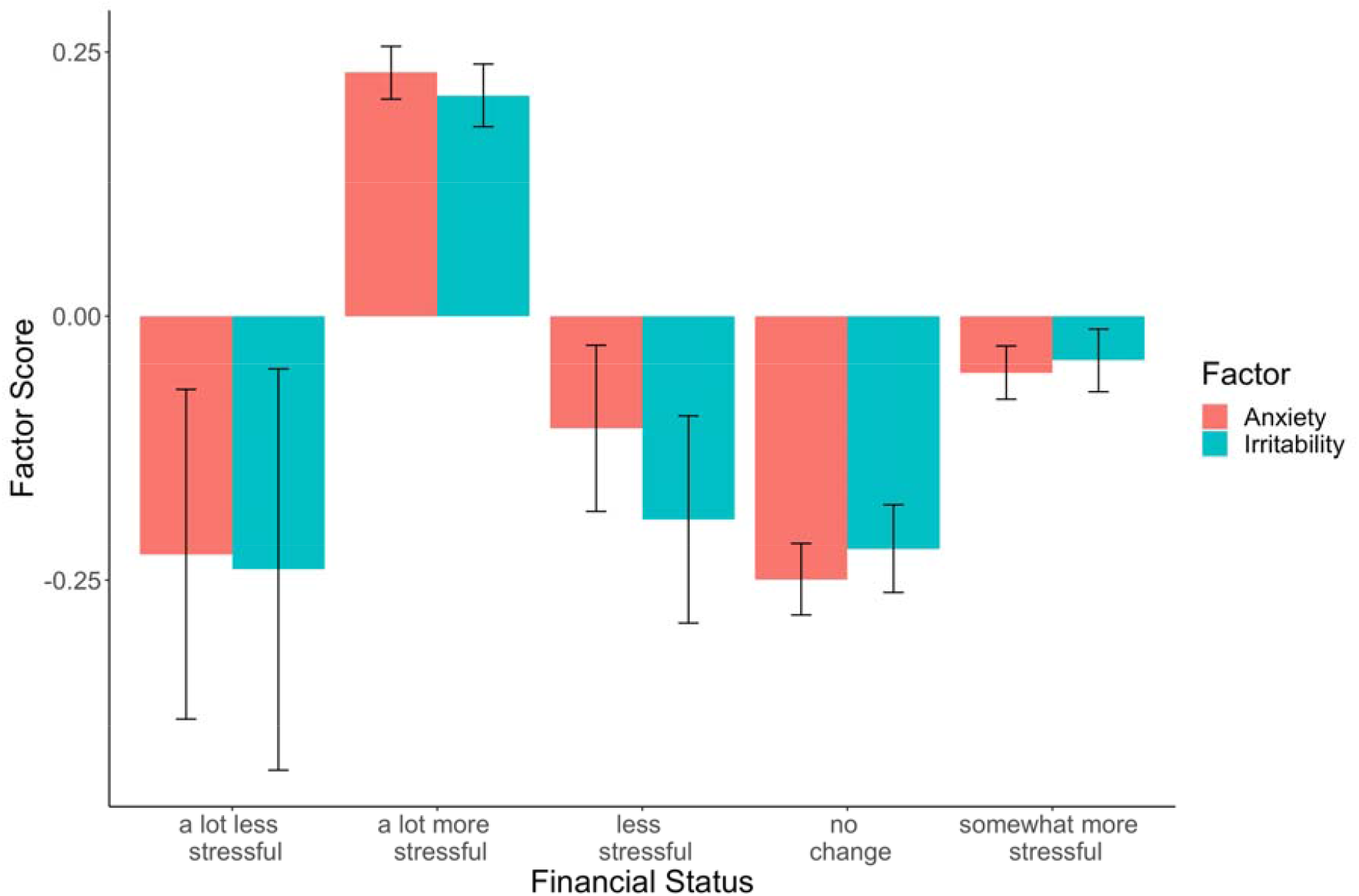
Mean Factor Scores for UNLV Students (n = 1,699) by Financial Status. Means and standard errors are presented.

**Figure 3:**
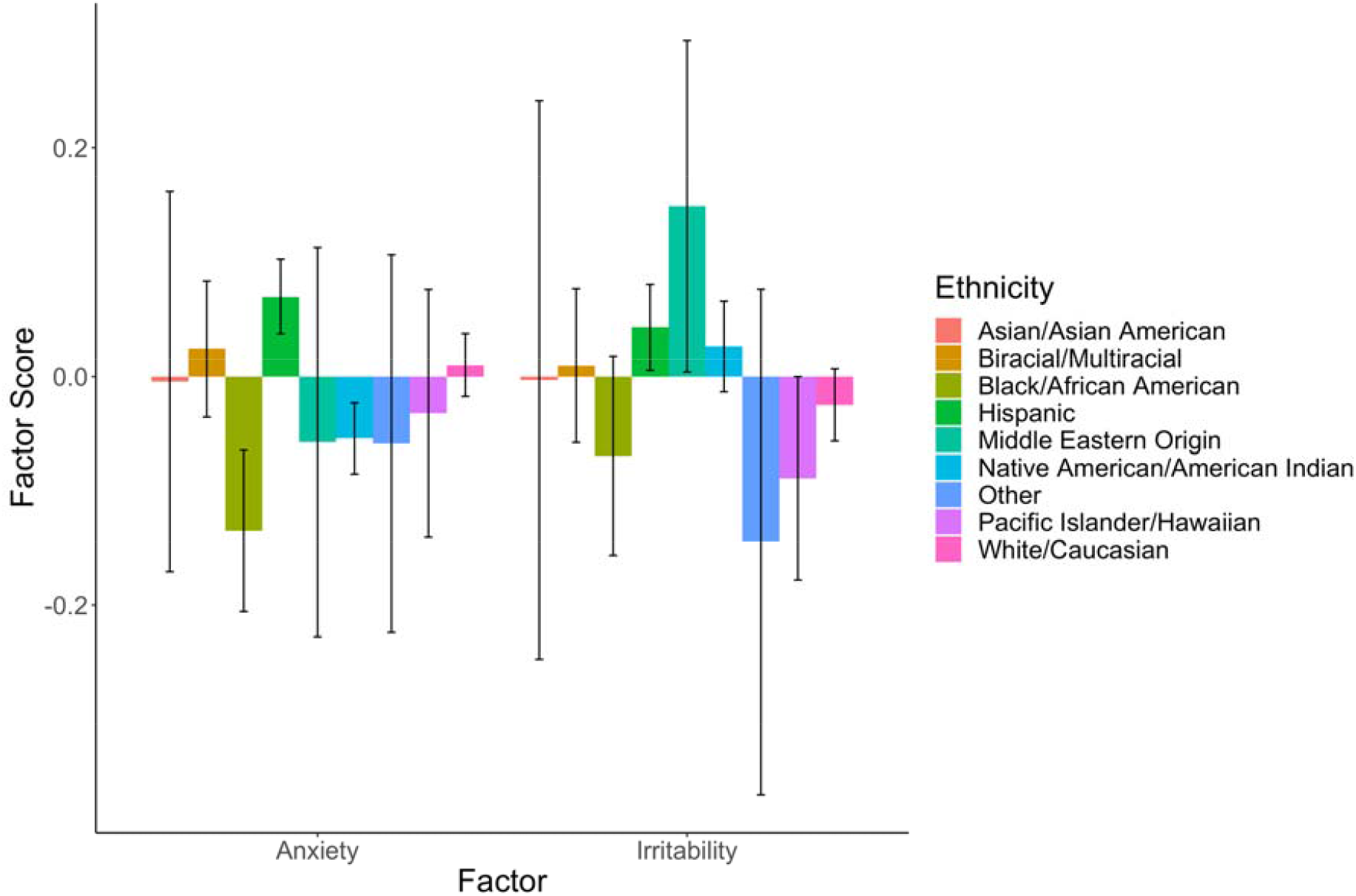
Mean Factor Scores for UNLV Students (n = 1,699) by Ethnicities. Means and standard errors are presented.

### Stepwise Linear Regression

The factor scores for all participants who completed PSS-10 were computed using a regression method. Stepwise linear regression with demographic variables (gender, age, financial situation, marital status, class standing, employment status, and ethnicity) as predictors was performed for two factors. Since most of the demographic variables are categorical, we created the dummy variables correspondingly.

Both male (β = -0.46, *P* < 0.01) and female (β = -0.41, *P* < 0.01) were highly associated with higher irritability **(Table 4)**. The students aged between 18-24 years old (β = 0.32, ***P*** < 0.01) and students aged between 25-34 years old (β = 0.27, ***P*** < 0.01) were also associated with higher irritability (**Table 4**). In addition, the students with a lot more stressful (β = 0.39, ***P*** < 0.01) and somehow more stressful financial status (β = 0.14, ***P*** < 0.01) are significantly associated with higher irritability. Ethnicities, employment status, and marital status of students were not found to be significantly associated with the factor “irritability” (**Table 4**).

**Table 4:**
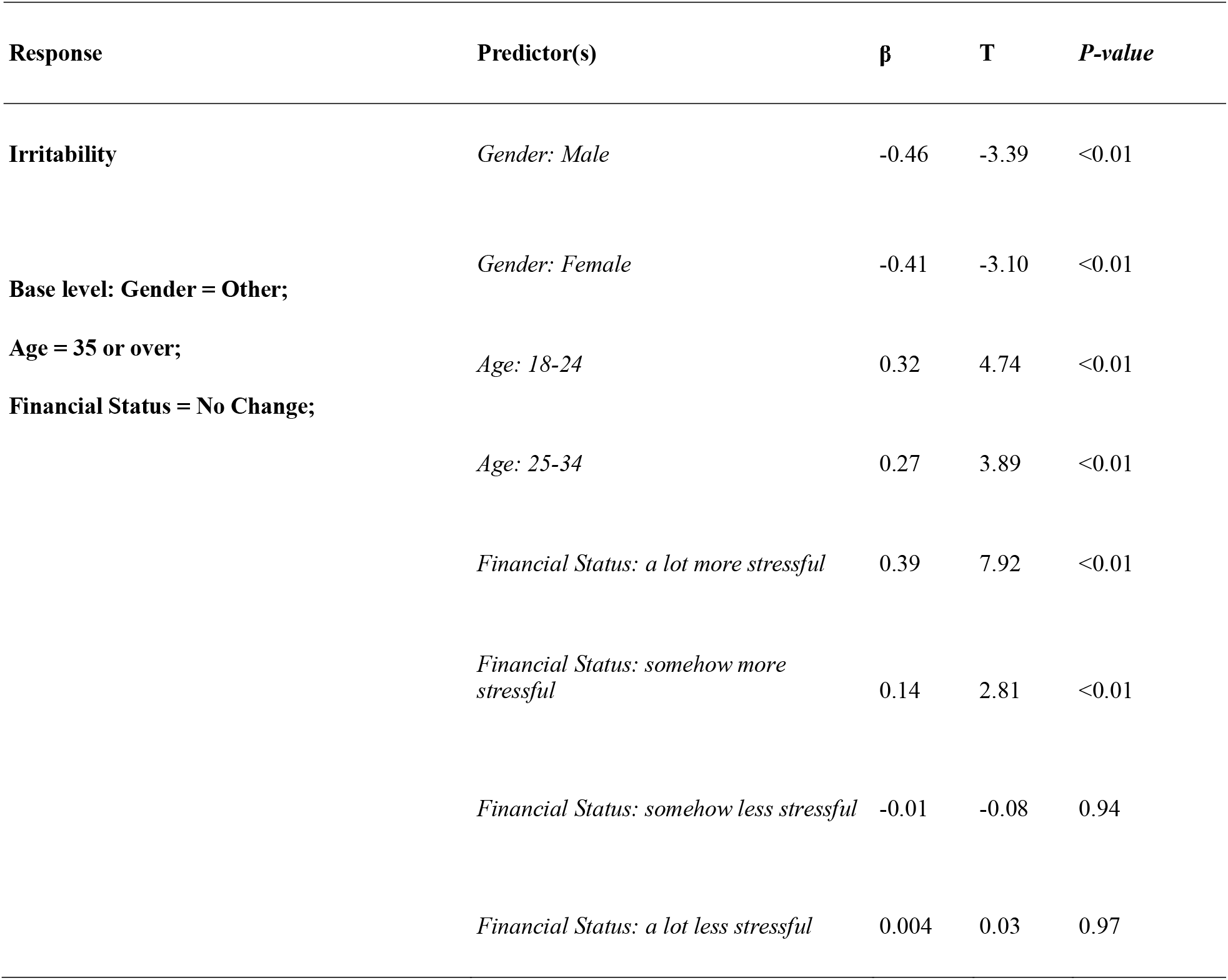
Results of Stepwise Linear Regression on “Irritability” with Categorical Predictors

Both male (β = -0.42, ***P*** < 0.01) and female (β = -0.32, ***P*** < 0.01) findings predicted anxiety (**Table 5**). While all other predictors were held as constants, females were associated with higher anxiety than males. Similar to the analysis on factor 1 (“Irritability”), the base level “other” was associated with higher anxiety in comparison to the other two groups (i.e., male and female). Stressful financial situations were associated with higher anxiety (“A lot more stressful”: β = 0.47, ***P*** < 0.01; “Somewhat more stressful”: β = 0.18, ***P*** < 0.01) (**Table 5**). In addition, the students aged between 18-24 years old (β = 0.15, ***P*** < 0.01) were also associated with higher anxiety (**Table 5**) compared to the students aged 45 or over. Ethnicities, employment status, and students’ marital status were not found to be significantly associated with the factor “anxiety” (**Table 5**).

**Table 5:**
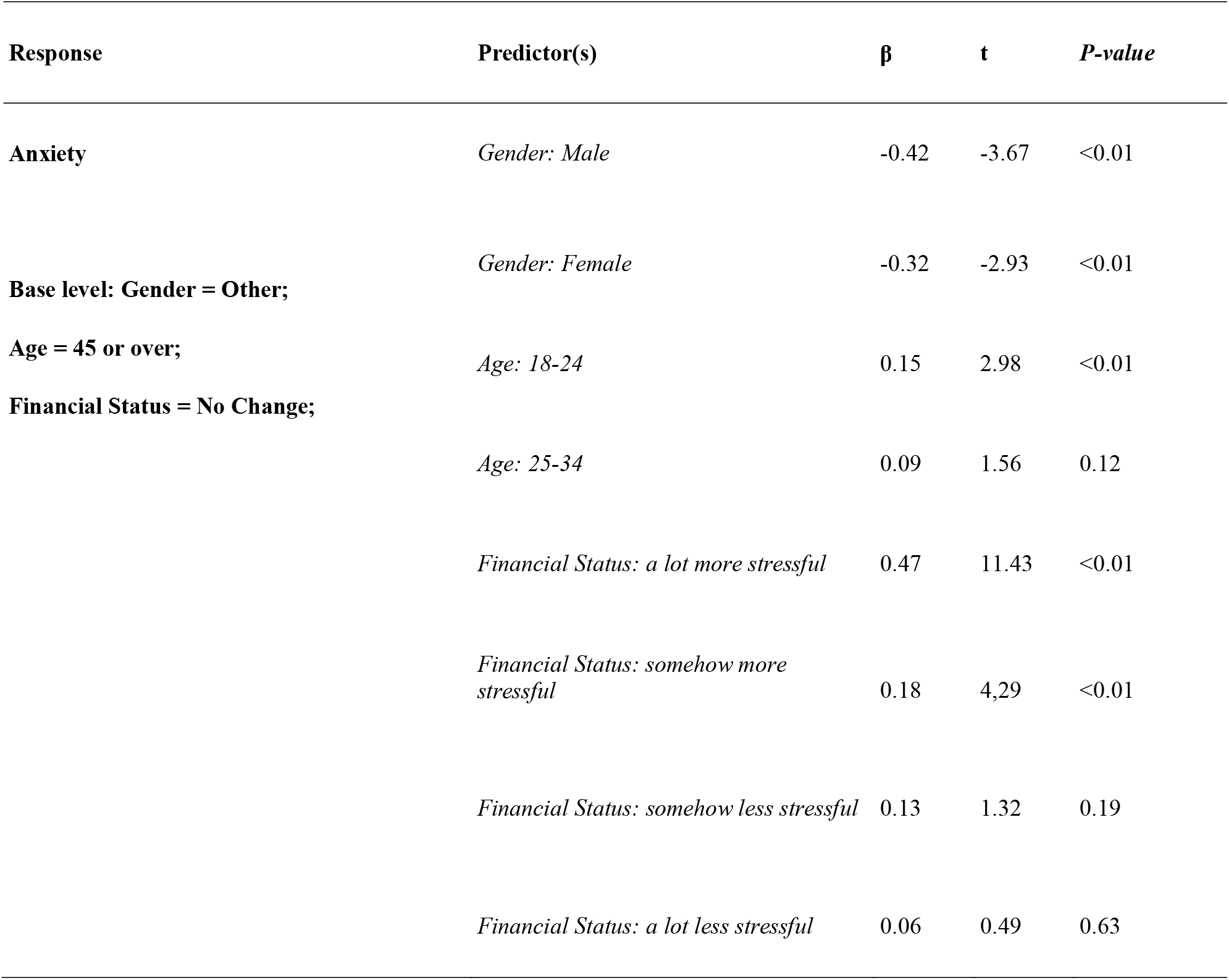
Results of Stepwise Linear Regression on “Anxiety” with Categorical Predictors

## Discussion

This study investigated how the COVID-19 pandemic has impacted the psychological status of college students and what demographic variables possibly contribute to such impacts. The factor analysis resulted in two factors: Anxiety and Irritability. The two-factor model is consistent with what was found by Wu and Amtmann ^24^. Furthermore, our stepwise regression on both factors revealed key risk factors for the prevalence of irritability and anxiety. Variables such as gender and financial situation demonstrated significant association with both factors in the analyses, while other variables were surprisingly insignificant such as ethnicity.

For the first factor, “Irritability,” students who identified themselves as ‘other’ gender scored significantly higher in this factor in comparison to the students who identified themselves as either male or female. This finding is reasonable since LGBTQ young persons might experience unique mental health problems compared to other gender groups during the COVID pandemic ^25^. However, female students are more likely to be irritable than male students in our study, which is consistent with the finding of Hou ^7^. Students who experience more stress financially are also associated with higher irritability. Among all the ethnic groups, the students with Middle East origin demonstrated higher irritability during the COVID pandemic than other ethnicities. However, as the stepwise linear regression suggests, the ethnicity of college students does not show up as a significant predictor of irritability. Our results also suggested that Ph.D. students and master’s students have lower irritability levels than others. This result might come as a surprising result since master’s students and Ph.D. students generally experience much pressure due to their coursework or research. However, due to their long-term experience coping with stress during their academic career, they might have developed an excellent ability to deal with stress under unexpected circumstances such as COVID. The students who were laid off during COVID and those who could not work due to disability generally have higher irritability than other groups, which is expected since the unemployment due to COVID led to mental health issues, as a previous study suggested ^26^.

The second factor, “Anxiety,” is associated with stressful financial status and gender, based on our stepwise regression analysis. Again, the students who identified themselves as “other” gender had significantly higher anxiety levels than males or females. Among all the ethnicities, the Hispanic-origin students had the highest level of anxiety. However, our stepwise linear regression suggests that students’ ethnicities are not significant predictors of anxiety. The widowed and separated students had higher anxiety levels than other students, which is expected since the loss of support from a loved one might reduce their ability to cope with stress. This result is also partially consistent with the finding of Nkire et al. ^27^

The factor analysis and stepwise linear regression confirm some observations found in the existing literature, such as the correlation of gender with anxiety and stress due to COVID. Interestingly, in contrast to what has been suggested by some researchers^6,12^, our results show that ethnicity seems not to be a significant risk factor in either irritability or anxiety. We speculate that the ethnically diverse UNLV student body and the overall social atmosphere that characterizes the university environment lead to less discrimination. Hence ethnicity is considered insignificant in predicting stress. We recommend that the university administration focuses on mental health policies that protect women and students who identify as other genders. We hope that this study raises awareness for all students’ growing mental health needs, with a priority to females, especially for students who identify as other genders. Furthermore, efforts should be made to provide low-cost mental health services for all students, particularly those who have lost their jobs and are in stressful financial situations due to the ongoing pandemic.

We realize that our survey study has several limitations. Although this study has a relatively large sample size with participants from diverse backgrounds, the study’s response rate is relatively low. In addition, compared to the student demographics data provided by UNLV during the 2020 fall semester, African Americans and Pacific Islanders have a lower proportion within our sample. This result indicates that African American and Pacific Islander students were under-represented in our sample, leading to potential volunteer bias. In addition, we only included the responses that fully completed the PSS-10 scales, which might lead to potential selection bias. Since most of the responses that did not complete the PSS-10 scales also did not complete the demographic questions, we could not check the differences between sample compositions of the responses that completed the PSS-10 scale and those that did not complete the PSS-10 scale.

## Conclusion

To our knowledge, this is the first study that assessed the perceived stress of university students in the US during the COVID-19 pandemic. We first investigated the relationship between each demographic variable and the perceived stress. Variables such as age, gender, marital status, employment status, financial status, and class standings are significantly associated with the perceived stress scores in our univariate analysis. Through exploratory factor analysis, we demonstrated that the PSS-10 scale could be summarized as a two-factor structure. After referring to previous studies, we identified two factors: “Irritability” and “Anxiety,” respectively. A stepwise regression analysis found that both factors are significantly associated with the gender of the participants and their financial status. However, no significant association was observed between both factors and ethnicity. This finding is consistent with the plot of mean factor scores for the ethnic groups. In summary, our findings will help identify students with increased risk for stress and mental health issues in pandemics of the Omicron variant and future crises.

## Supporting information

Supplemental File 1: PSS-10 Scale

## Data Availability

All data produced in the present study are available upon reasonable request to the authors.

## Acknowledgements

We would like to thank all the student participants, the Office of Registrar, the Institutional Review Board, the Office of Information Technology, and the Department of Epidemiology and Biostatistics at the University of Nevada, Las Vegas.

## Author Contributions

Conceptualization, BL, EH, CL and QW; methodology, BL, EH and CL; software, BL..; validation, BL, EH, CL and QW; formal analysis, BL; investigation, BL, EH, CL and QW; resources, BL, EH, CL and QW; data curation, BL, EH, CL and QW; writing—original draft preparation, BL, EH and CL; writing—review and editing, BL, EH, CL and QW; visualization, BL, EH and CL; supervision, QW; project administration, QW

## Funding

This research received no external funding. Competing Interest: None declared.

## Ethics Approval

The study was conducted according to the guidelines of the Declaration of Helsinki and approved by the Institutional Review Board of the University of Nevada, Las Vegas (approval number 1632208, effective on November 3rd, 2020).

## Participants’ Consent for Publication

Written informed consent has been obtained from the participants in order to publish this paper.

## Provenance and Peer Review

Not commissioned; externally peer-reviewed

## Data Availability Statement

All data relevant to the study are available upon request.

## Conflicts of Interest

The authors declare no conflict of interest.

## Notes

### Competing Interest Statement

The authors have declared no competing interest.

### Funding Statement

This study did not receive any funding.

### Author Declarations

IRB of University of Nevada, Las Vegas gave ethical approval for this work.

