## Supplemental File 1: PSS-10 Scale for "Impact of COVID-19 on College Students’ at One of the Most Diverse Campuses in the United States: A Factor Analysis of Survey Data"

### Perceived Stress Scale (PSS-10)

The questions in this scale ask you about your feelings and thoughts **during the last month**.

In each case, you will be asked to indicate by circling how often you felt or thought a certain way.

**0 = Never 1 = Almost Never 2 = Sometimes 3 = Fairly Often 4 = Very Often**

| | | | | | |
|---|---|---|---|---|---|
| 1. In the last month, how often have you been upset because of something that happened unexpectedly? | 0 | 1 | 2 | 3 | 4 |
| 2. In the last month, how often have you felt that you were unable to control the important things in your life? | 0 | 1 | 2 | 3 | 4 |
| 3. In the last month, how often have you felt nervous and “stressed”? | 0 | 1 | 2 | 3 | 4 |
| 4. In the last month, how often have you felt confident about your ability to handle your personal problems? | 0 | 1 | 2 | 3 | 4 |
| 5. In the last month, how often have you felt that things were going your way? | 0 | 1 | 2 | 3 | 4 |
| 6. In the last month, how often have you found that you could not cope with all the things that you had to do? | 0 | 1 | 2 | 3 | 4 |
| 7. In the last month, how often have you been able to control irritations in your life? | 0 | 1 | 2 | 3 | 4 |
| 8. In the last month, how often have you felt that you were on top of things? | 0 | 1 | 2 | 3 | 4 |
| 9. In the last month, how often have you been angered because of things that were outside of your control? | 0 | 1 | 2 | 3 | 4 |
| 10. In the last month, how often have you felt difficulties were piling up so high that you could not overcome them? | 0 | 1 | 2 | 3 | 4 |
