## Supplemental File 2: Supplementary Figures for "Impact of COVID-19 on College Students’ at One of the Most Diverse Campuses in the United States: A Factor Analysis of Survey Data"

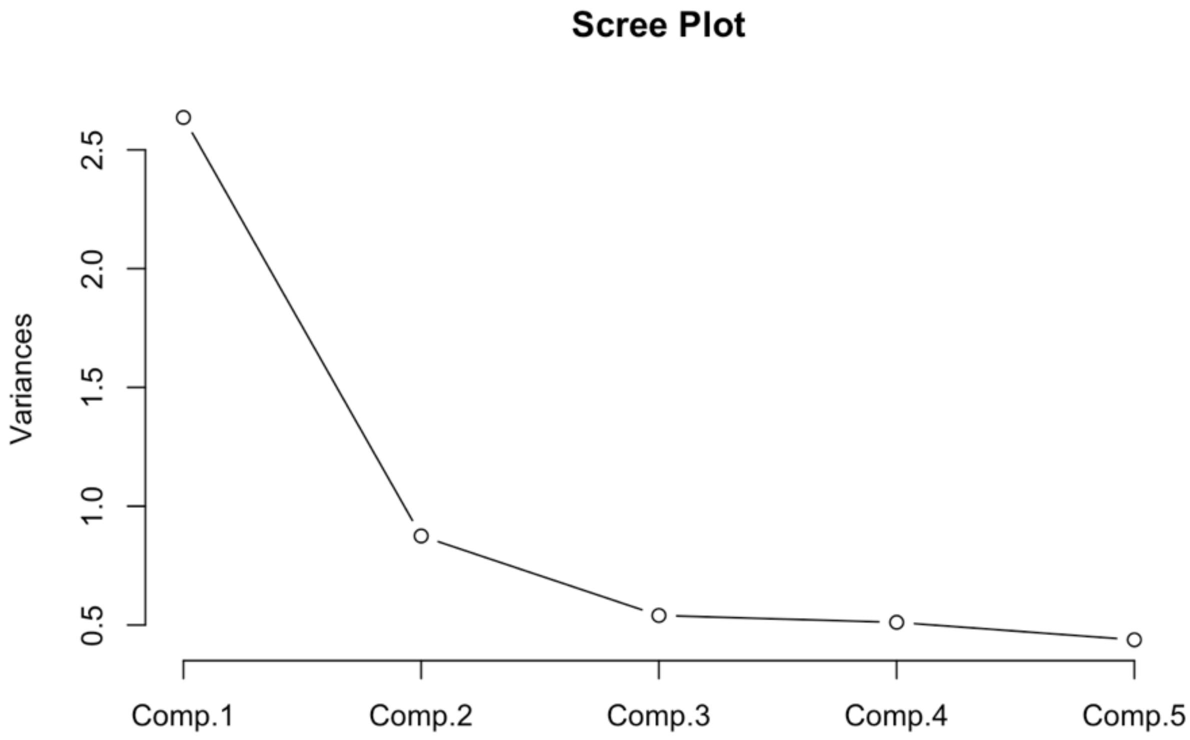


**Supplementary Figure 1:** Scree Plot for Factor Analysis with PSS-10 Scale

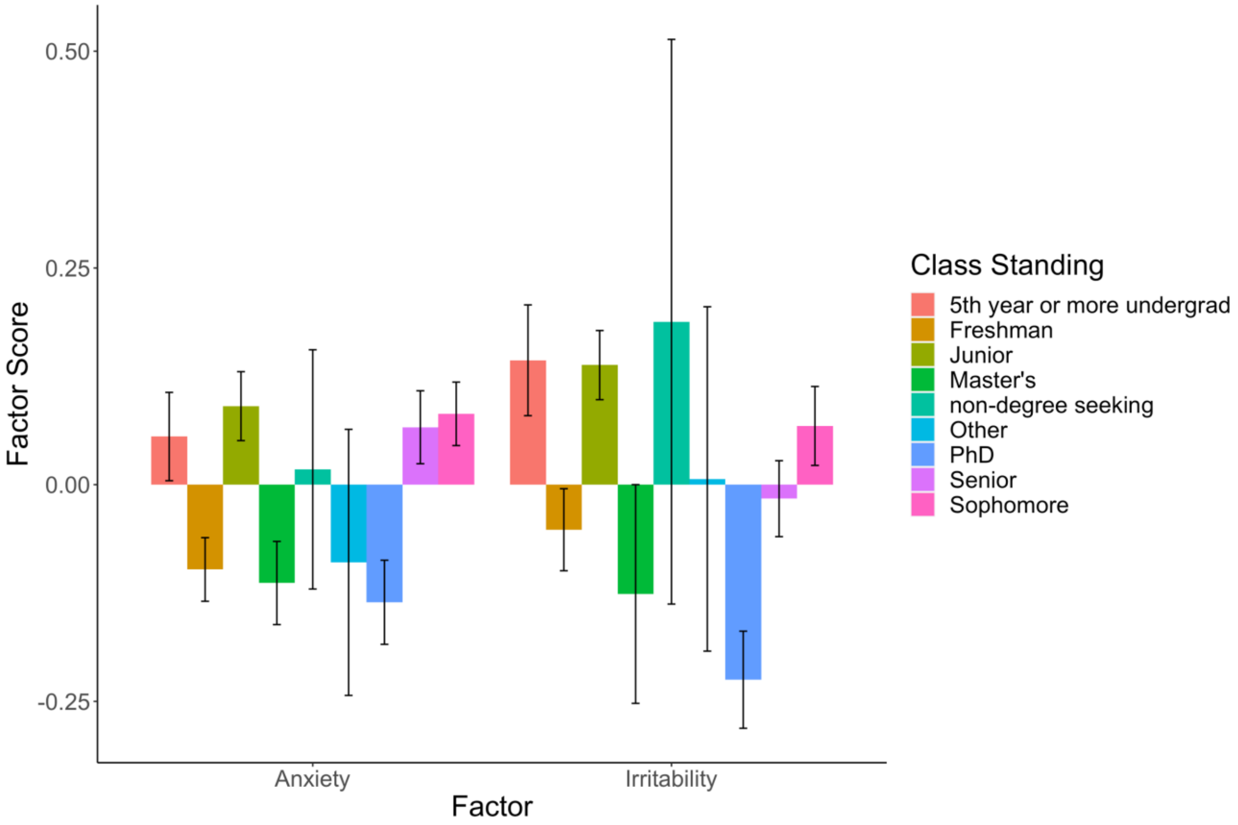


The colored bars represent the mean factor scores and the error bars are presented with all colored bars.

**Supplementary Figure 2:** Mean Factor Scores for UNLV Students (n = 1,699) by Class Standing.

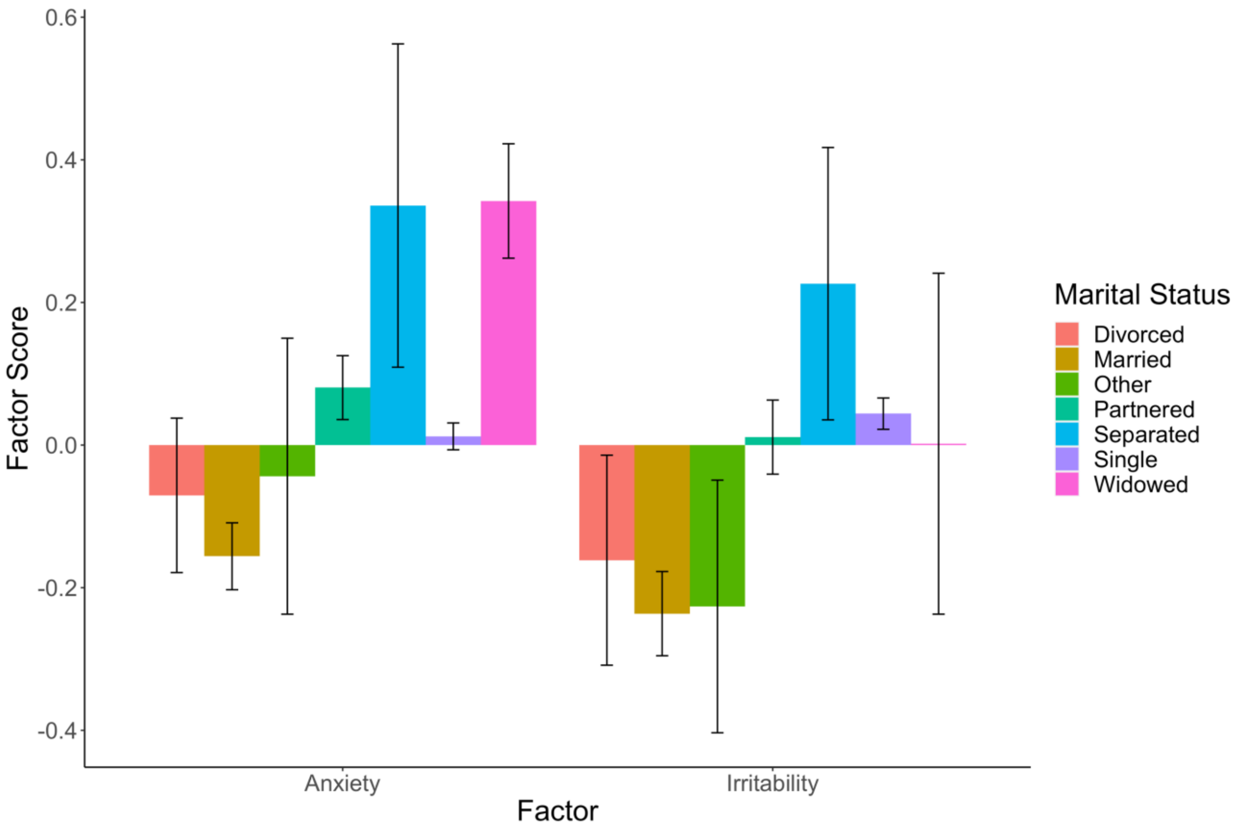


The colored bars represent the mean factor scores and the error bars are presented with all colored bars.

**Supplementary Figure 3:** Mean Factor Scores for UNLV Students (n = 1,699) by Marital Status
